# AI-enabled CT-guided end-to-end quantification of total cardiac activity in 18FDG cardiac PET/CT for detection of cardiac sarcoidosis

**DOI:** 10.1101/2024.09.20.24314081

**Authors:** Robert JH Miller, Aakash Shanbhag, Anna M Marcinkiewicz, Helen Struble, Hidesato Fujito, Evan Kransdorf, Paul Kavanagh, Joanna X. Liang, Valerie Builoff, Damini Dey, Daniel S Berman, Piotr J Slomka

## Abstract

**Purpose:** [18F]-fluorodeoxyglucose ([18F]FDG) positron emission tomography (PET) plays a central role in diagnosing and managing cardiac sarcoidosis. We propose a fully automated pipeline for quantification of [18F]FDG PET activity using deep learning (DL) segmentation of cardiac chambers on computed tomography (CT) attenuation maps and evaluate several quantitative approaches based on this framework.

**Methods:** We included consecutive patients undergoing [18F]FDG PET/CT for suspected cardiac sarcoidosis. DL segmented left atrium, left ventricular(LV), right atrium, right ventricle, aorta, LV myocardium, and lungs from CT attenuation scans. CT-defined anatomical regions were applied to [18F]FDG PET images automatically to target to background ratio (TBR), volume of inflammation (VOI) and cardiometabolic activity (CMA) using full sized and shrunk segmentations.

**Results:** A total of 69 patients were included, with mean age of 56.1 ± 13.4 and cardiac sarcoidosis present in 29 (42%). CMA had the highest prediction performance (area under the receiver operating characteristic curve [AUC] 0.919, 95% confidence interval [CI] 0.858 – 0.980) followed by VOI (AUC 0.903, 95% CI 0.834 – 0.971), TBR (AUC 0.891, 95% CI 0.819 – 0.964), and maximum standardized uptake value (AUC 0.812, 95% CI 0.701 – 0.923). Abnormal CMA (≥1) had a sensitivity of 100% and specificity 65% for cardiac sarcoidosis. Lung quantification was able to identify patients with pulmonary abnormalities.

**Conclusion:** We demonstrate that fully automated volumetric quantification of [18F]FDG PET for cardiac sarcoidosis based on CT attenuation map-derived volumetry is feasible, rapid, and has high prediction performance. This approach provides objective measurements of cardiac inflammation with consistent definition of myocardium and background region.

## INTRODUCTION

Cardiac sarcoidosis is an inflammatory cardiomyopathy characterized by the presence of non-caseating granulomas [1]. The disorder leads to heart failure, ventricular and atrial arrhythmias, and conduction system disease [1]. Accurate diagnosis is central to targeting immunosuppressive medications which can decrease myocardial inflammation, and therefore decrease the incidence of adverse events [1–3]. However, diagnosis of cardiac sarcoidosis can be challenging and relies on a combination of clinical and imaging criteria [4, 5]. Cardiac positron emission tomography (PET) with [18F]-fluorodeoxyglucose ([18F]FDG) plays a central role in establishing the diagnosis by identifying myocardial inflammation [4–8].

While PET imaging is widely used to aid in the diagnosis of sarcoidosis, visual identification of abnormal myocardial hypermetabolism requires significant expertise and is inherently subjective, which may be particularly difficult in cases with borderline myocardial uptake. Additionally, it does not provide treating clinicians with detailed information regarding the severity of inflammation. Quantitative analysis of cardiac hypermetabolism can be performed using maximum standardized uptake value (SUVmax) or volumetric measurements (which capture both volume and intensity of [18F]FDG uptake). Patients with more extensive cardiac inflammation are more likely to have cardiac sarcoidosis [9], but are also more likely to benefit from immunosuppression [10]. We previously demonstrated that volume of involvement (VOI) and cardiometabolic activity (CMA), quantified from [18F]FDG PET studies, had high diagnostic accuracy for cardiac sarcoidosis [11]. However, these measurements required manual annotations, which take about 10 minutes per study, and are prone to interobserver variability related to placement of both the background region and region of interest. Additionally, there remains uncertainty regarding the optimal method for quantifying myocardial inflammation [8].

We recently demonstrated that cardiac chambers can be segmented accurately from non-contrast computed tomography (CT) attenuation correction (CTAC) scans [12], utilizing a foundational deep learning CT segmentation model [13]. This approach can be translated to “hot spot” imaging such as [18F]FDG PET, to provide fully automated quantification of cardiac activity in selected cardiac chambers. Importantly, this method allows for rapid and consistent evaluation of multiple structures simultaneously, facilitating automatic placement of background regions and rapid comparison between quantitative approaches and alternative region placements. We implemented and evaluated the feasibility and diagnostic accuracy of this approach in patients undergoing [18F]FDG PET to determine the optimal approach for image quantification.

## MATERIALS AND METHODS

### Study Population

For this study, we utilized our previously described cohort of patients with suspected cardiac sarcoidosis [11]. In brief, we included consecutive patients undergoing [18F]FDG PET for suspected cardiac sarcoidosis. Patients who were imaged without adequate dietary preparation (n=1) were excluded. In patients with multiple studies, only the first study was included (n=29). A total of 69 patients were included in the analysis. This study was approved by the institutional review board at Cedars-Sinai Medical Center including waiver of informed consent for use of retrospective data. Analyses were performed on de-identified image sets consistent with Health Insurance Portability and Accountability Act compliance.

### Demographic Information

Demographic information including age, sex, height, weight, body mass index and medical history were collected prospectively at the time of PET imaging. Diagnosis of cardiac sarcoidosis was determined retrospectively after reviewing all available clinical, imaging, and pathologic information. Five patients underwent cardiac transplant and diagnosis was based on pathologic examination of the explanted heart (4 with cardiac sarcoidosis, 1 without). An additional two patients had cardiac sarcoidosis diagnosed on the basis of positive endomyocardial biopsy. The remaining patients were categorized as having definite or probable cardiac sarcoidosis using the Japanese Ministry of Health and Welfare criteria [4], which includes regional [18F]FDG uptake on PET [14]. Patients with definite or probable cardiac sarcoidosis were considered to have cardiac sarcoidosis.

### PET Imaging Acquisition

Images were acquired with a whole-body PET/CT scanner (Biograph-64, Siemens, Munich, Germany). All patients were instructed to eat a high-fat/low-carbohydrate diet for 36 hours prior to the scan, with fasting for at least 8 hours prior to the scan. Patients who did not comply with the dietary protocol were re-scheduled for imaging when dietary compliance could be ensured. Scans were not performed if fasting blood glucose was >200 mg/dL. [18F]FDG imaging was performed approximately 60 minutes after injection of 5-15 mCi of [18F]FDG (based on patient weight) and acquired over 15-20 minutes [15].

CTAC scans were acquired with normal breathing, without ECG gating, spiral mode with pitch 1.5, tube voltage 100 kVp, tube current–time product 11–13 mAs, and a full field-of-view. Reconstruction was performed with a slice thickness of 3mm. Hybrid image sets were evaluated for significant misregistration during processing, and if misregistration was noted during clinical reconstruction the data were re-reconstructed after adjustment, according to standard clinical protocols.

### Image Analysis

Semi-quantitative assessments of resting perfusion were assessed using summed rest score (SRS) with dedicated software (QPET, Cedars-Sinai Medical Center, Los Angeles, California) [16, 17]. Established thresholds for abnormal perfusion (SRS ≥3) were used as the criteria for perfusion deficit [17]. Myocardial hypermetabolism was assessed visually by an experienced cardiologists at the time of clinical reporting. Visual interpretation was used as the criteria for positive PET.

### CTAC Heart Segmentation

A previously validated deep learning model (TotalSegmentator), was used to segment structures from CTAC [13]. In brief, the model utilizes a no new-net (nnU-Net) architecture (which automatically optimized hyperparameters) to automatically segment anatomic structures from CT images [13]. During model development the lungs could be segmented directly from expert annotation of non-contrast images, with Dice scores >0.98 in the testing set [13]. For cardiac structures, the model was trained using expert annotations from contrast images which were transferred to registered non-contrast images. We recently demonstrated excellent correlations in an external population between volumes manually segmented from gated, contrast-enhanced CT and automated segmentations from non-contrast, ungated CT (left atrium [LA] volume Spearman r=0.926, left ventricle [LV] myocardium Spearman r=0.947, LV volume Spearman r=0.793, right atrial [RA] volume Spearman r=0.893, and right ventricular (RV) volume Spearman r=0.922) [12].

### PET Lung Quantification

To further evaluate the potential utility of applying CT-based segmentation and quantification, we also evaluated methods for assessing PET lung quantification. We utilized lung segmentations from the TotalSegmentator [13], then quantified lung activity within both lungs. We evaluated SUVmax, as well as target to background ratio (TBR) based on maximal activity relative to mean activity in the lungs as background. Since no standardized thresholds for abnormal activity exist, we utilized mean + 2 standard deviations as the threshold for abnormal lung activity. Lung activity above background was measured using both volume above threshold (lung metabolic volume [LMV]) and activity above background (lung metabolic activity [LMA]). We evaluated prediction performance for pulmonary abnormalities (lymphadenopathy, pulmonary nodules, or consolidations) based on expert interpretation of CT radiographic findings.

### PET Cardiac Quantification

The LA, LV, RA, RV and aorta segmentations were used as background regions for cardiac hypermetabolism quantification. The LV myocardial volume was used as the region of interest for cardiac quantification. For background activity, we evaluated full sized segmentations as well as segmentations that were volumetrically reduced by 50% to avoid partial volume effects. These “shrunk” segmentations were centered within the original segmentation. In total, we evaluated 6 potential region definitions (LA-full, LA-shrunk, RA-full, RA-shrunk, Aorta-full, Aorta-shrunk) for computation of the background activity. We applied four potential thresholds for abnormal activity (max counts, 1.3 * max counts, 1.5* max counts, and mean counts + 2 standard deviations) based on previous work [9, 11]. These segmentations were then translated to co-registered [18F]FDG PET images. An overview of the segmentation methods and overall workflow for cardiac quantification are shown in **Figure 1**. In our primary analyses, we utilized maximum background counts as the threshold for abnormal activity.

**Figure 1.**
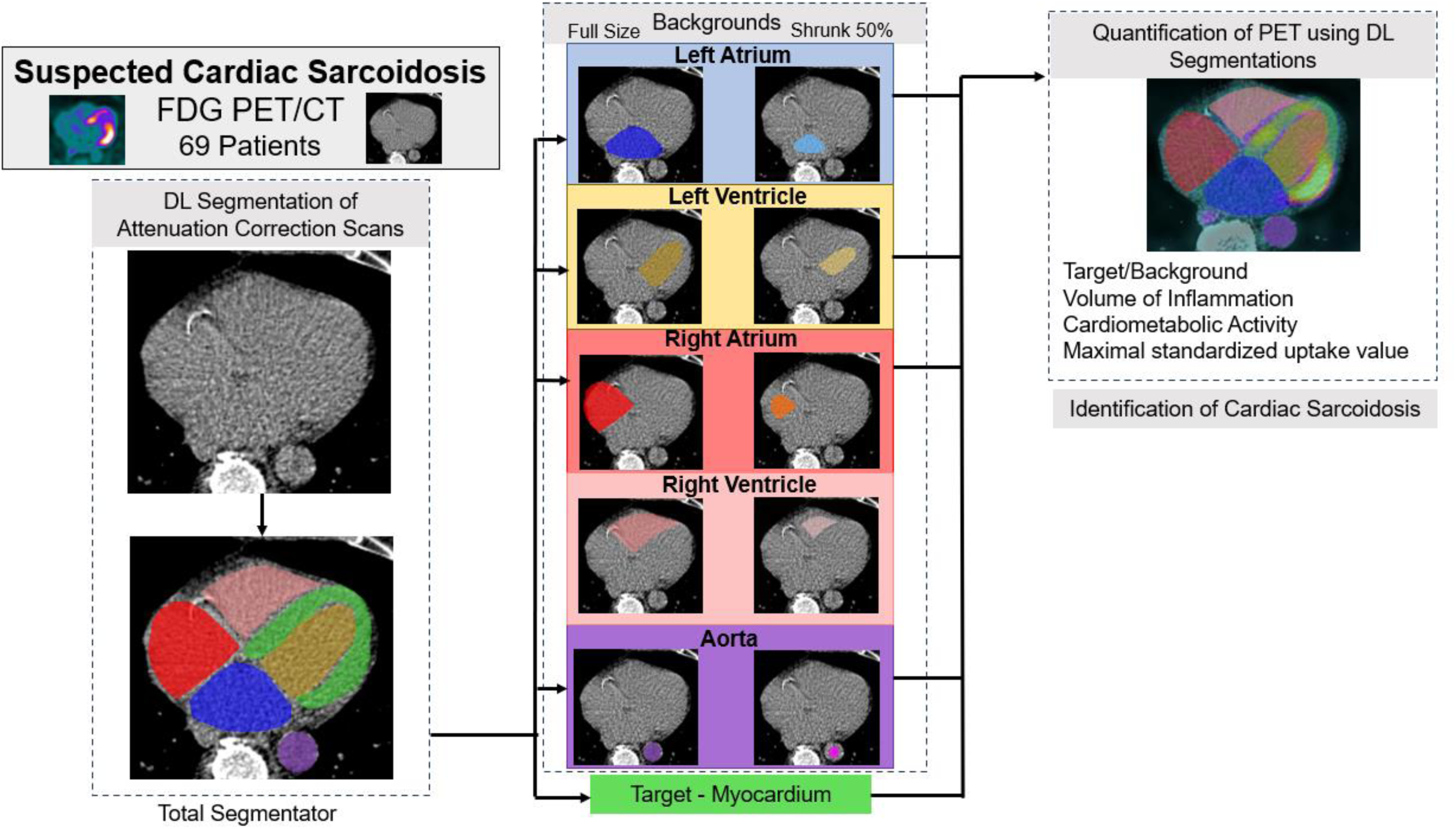
Patients undergoing [18F]-fluorodeoxyglucose ([18F]FDG) positron emission tomography (PET) for suspected cardiac sarcoidosis were included. Deep learning (DL) segmented cardiac structures from associated computed tomography (CT) images, were used to define background and target regions. Those segmentations were then applied to quantify PET activity, demonstrating excellent prediction for cardiac sarcoidosis.

SUVmax was quantified as maximum uptake within the LV myocardial volume. TBR, VOI, and CMA were computed utilizing 10 different background definitions. TBR was calculated as SUVmax for LV myocardium divided by mean background activity. VOI is calculated as the volume of myocardium above threshold for abnormal. CMA is calculated as the total activity within the LV myocardium above the threshold for abnormal.

### Statistical Analysis

Continuous variables were summarized as mean +/- standard deviation and compared with a Student’s t-test or Mann-Whitney U test as appropriate. Categorical variables were summarized as number (proportion) and compared using a chi-square or Fisher exact test as appropriate. Receiver operating characteristic (ROC) curves were generated to assess prediction performance for cardiac sarcoidosis. Area under the ROC curve (AUC) was compared using the method established by DeLong et al. [18]. Optimal thresholds were determined using the Youden index. All statistical tests were two-sided and a p-value <0.05 was considered significant, with analyses performed in Stata version 13 (StataCorp, College Station, Texas).

## RESULTS

### Patient Population

A total of 69 patients were included, with mean age 56.1 ± 13.4 years and 41 (59.4%) male patients. Cardiac sarcoidosis was present in 29 (42%) patients. Patient characteristics stratified by the presence of cardiac sarcoidosis are shown in **Table 1**. There was no significant difference in median age between patients with cardiac sarcoidosis compared to those without cardiac sarcoidosis (51 vs 60, p=0.175). More patients with cardiac sarcoidosis had atrioventricular block (41% vs 13%, p=0.010) and reduced left ventricular function (86% vs 35%, p<0.001).

**Table 1:** Clinical characteristics stratified by diagnosis of cardiac sarcoidosis. AV – atrioventricular, IQR – interquartile range, LVEF – left ventricular ejection fraction, PET – positron emission tomography.

|  | No Cardiac Sarcoidosis<br>N=40 | Cardiac Sarcoidosis<br>N=29 | p-value |
| --- | --- | --- | --- |
| Age, median (IQR) | 60 (46, 68) | 51 (45, 59) | 0.175 |
| Male, n (%) | 22 (55%) | 19 (66%) | 0.460 |
| Cardiac Sarcoidosis Features |  |  |  |
| Advanced AV block | 5 (13%) | 12 (41%) | 0.010 |
| Basal Septal Thinning | 0 (0%) | 3 (10%) | 0.071 |
| Visually Abnormal PET | 2 (5%) | 24 (83%) | <0.001 |
| Decreased LVEF | 14 (35%) | 25 (86%) | <0.001 |
| Abnormal electrocardiogram | 15 (38%) | 14 (48%) | 0.461 |
| Abnormal echocardiogram | 1 (3%) | 1 (3%) | 1.000 |
| Perfusion deficit | 8 (20%) | 17 (59%) | 0.002 |
| Late gadolinium enhancement | 14 (35%) | 8 (28%) | 0.605 |

Fully automated quantification was performed in ~16 seconds per patient. Imaging characteristics stratified by the presence of cardiac sarcoidosis are summarized in **Table 2**. Patients with cardiac sarcoidosis had higher values for all quantitative metrics (all p<0.01).

**Table 2:** Imaging characteristics stratified by diagnosis of cardiac sarcoidosis. CMA – cardiometabolic activity, IQR – interquartile range, SUVmax – maximum standardized uptake value, VOI – volume of inflammation.

|  | No Cardiac Sarcoidosis<br>N=40 | Cardiac Sarcoidosis<br>N=29 | p-value |
| --- | --- | --- | --- |
| SUVmax, median (IQR) | 2.23 (1.89, 2.62) | 4.34 (2.54, 8.80) | <0.001 |
| Full left atrium, median (IQR) |  |  |  |
| Target to Background Ratio | 1.30 (1.22, 1.49) | 2.11 (1.75, 4.98) | <0.001 |
| VOI | 0.0 (0.0, 0.6) | 7.2 (0.1, 51.7) | <0.001 |
| CMA | 0.0 (0.0, 1.3) | 35.7 (0.2, 224) | <0.001 |
| Shrunk left atrium, median (IQR) |  |  |  |
| Target to Background Ratio | 1.28 (1.18, 1.47) | 2.18 (1.73, 5.23) | <0.001 |
| VOI | 0.0 (0.0, 1.2) | 17.5 (3.3, 49.0) | <0.001 |
| CMA | 0.0 (0.0, 5.1) | 98.4 (28.7, 303) | <0.001 |
| Full left ventricle, median (IQR) |  |  |  |
| Target to Background Ratio | 1.42 (1.30, 1.63) | 2.16 (1.72, 4.11) | <0.001 |
| VOI | 0.0 (0.0, 0.1) | 0.2 (0.0, 1.6) | 0.009 |
| CMA | 0.0 (0.0, 0.2) | 0.6 (0.0, 2.3) | 0.006 |
| Shrunk left ventricle, median (IQR) |  |  |  |
| Target to Background Ratio | 1.42 (1.30, 1.62) | 2.16 (1.71, 4.11) | <0.001 |
| VOI | 0.0 (0.0, 1.6) | 3.8 (0.8, 9.7) | <0.001 |
| CMA | 0.1 (0.0, 3.5) | 116 (2.5, 41.9) | <0.001 |
| Full right atrium, median (IQR) |  |  |  |
| Target to Background Ratio | 1.35 (1.26, 1.46) | 2.07 (1.83, 4.75) | <0.001 |
| VOI | 0.0 (0.0, 0.0) | 13.2 (2.4, 42.5) | <0.001 |
| CMA | 0.0 (0.0, 0.1) | 37 (9.9, 216) | <0.001 |
| Shrunk right atrium, median (IQR) |  |  |  |
| Target to Background Ratio | 1.27 (1.20, 1.44) | 2.02 (1.74, 4.83) | <0.001 |
| VOI | 0.0 (0.0, 0.4) | 37.7 (3.6, 92.6) | <0.001 |
| CMA | 0.0 (0.0, 0.9) | 92.2 (20.2, 289) | <0.001 |
| Full right ventricle, median (IQR) |  |  |  |
| Target to Background Ratio | 1.45 (1.35, 1.63) | 2.10 (1.90, 4.27) | <0.001 |
| VOI | 0.0 (0.0, 0.1) | 3.0 (0.2, 7.1) | <0.001 |
| CMA | 0.0 (0.0, 0.1) | 14.0 (1.3, 27.3) | <0.001 |
| Shrunk right ventricle, median (IQR) |  |  |  |
| Target to Background Ratio | 1.35 (1.24, 1.51) | 2.08 (1.79, 4.28) | <0.001 |
| VOI | 0.0 (0.0, 0.3) | 11.0 (3.3, 20.5) | <0.001 |
| CMA | 0.0 (0.0, 0.5) | 31.1 (11.5, 94.1) | <0.001 |
| Full Aorta, median (IQR) |  |  |  |
| Target to Background Ratio | 1.17 (1.08, 1.35) | 2.16 (1.51, 4.64) | <0.001 |
| VOI | 0.0 (0.0, 0.0) | 0.1 (0.0, 37.6) | <0.001 |
| CMA | 0.0 (0.0, 0.0) | 0.1 (0.0, 169) | <0.001 |
| Shrunk Aorta, median (IQR) |  |  |  |
| Target to Background Ratio | 1.12 (1.07, 1.30) | 1.92 (1.57, 4.64) | <0.001 |
| VOI | 0.0 (0.0, 0.0) | 11.36 (0, 59.6) | <0.001 |
| CMA | 0.0 (0.0, 0.0) | 48.1 (0.0, 181) | <0.001 |

### Prediction Performance for Cardiac Sarcoidosis

Prediction performance for cardiac sarcoidosis using CMA is shown in **Figure 2**. LA-shrunk (AUC 0.919, 95% confidence interval [CI] 0.858 – 0.980) had the highest prediction performance, and was significantly better compared to RA-full (AUC 0.837, 95% CI 0.739 – 0.936), LA-full (AUC 0.787, 95% CI 0.679 – 0.895), either LV background (full: AUC 0.688, 95% CI 0.564 – 0.812; shrunk: AUC 0.805, 95% CI 0.699 – 0.911), and either aorta background (full: AUC 0.752, 95% CI 0.656 – 0.847; shrunk: AUC 0.796, 95% CI 0.695 – 0.896) (all p<0.05). For comparison, the AUC for visual interpretation (which is included in the criteria for diagnosis) was 0.872 (95% CI 0.789 – 0.954). Similar results were seen for the prediction performance of VOI (**Supplemental Figure 1**). There were no significant differences in prediction performance using TBR (**Supplemental Figure 2**).

**Figure 2.**
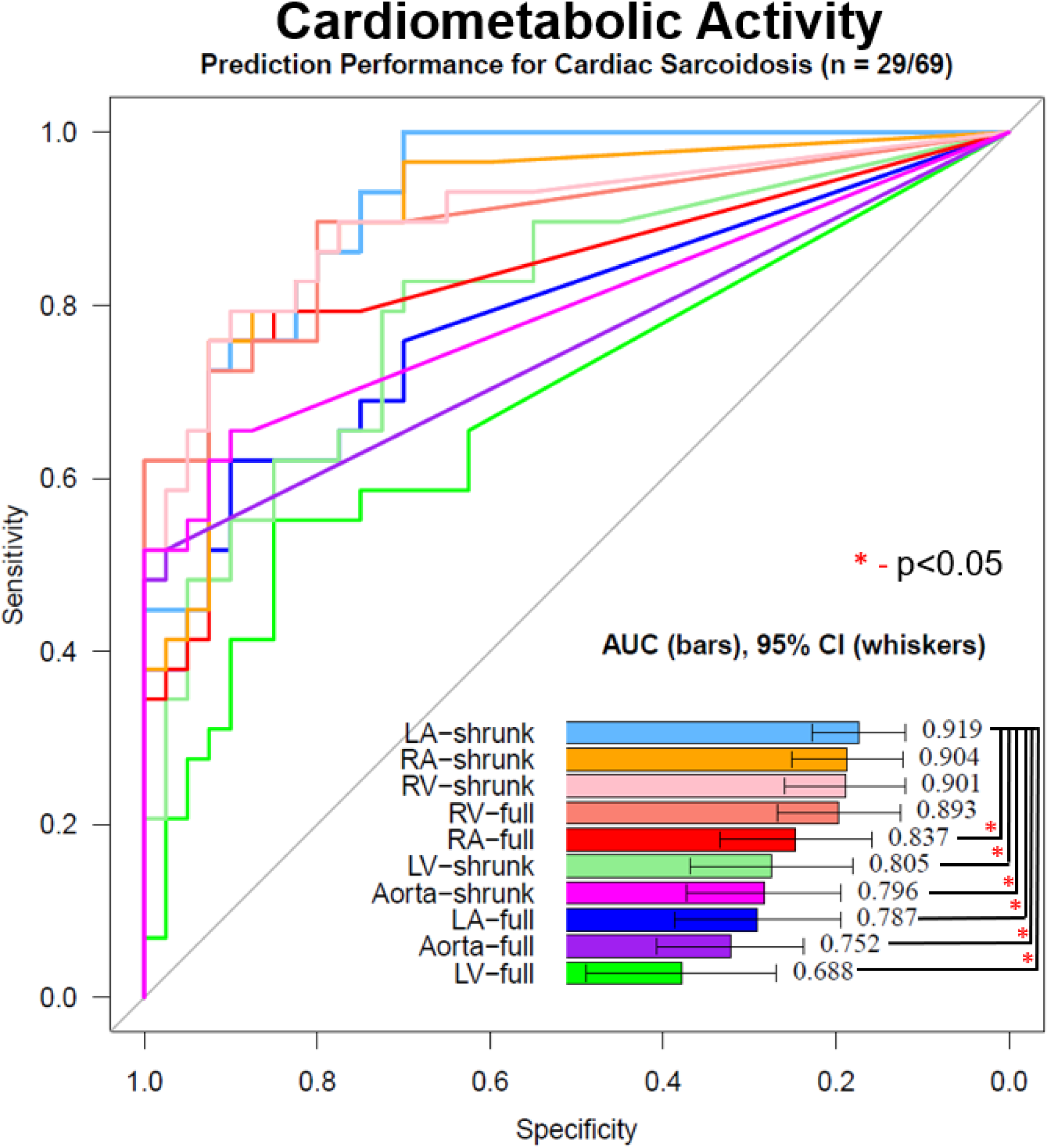
Prediction performance for cardiac sarcoidosis using different methods for quantifying cardiometabolic activity. Segments were derived using deep learning from computed tomography attenuation imaging. Cardiac sarcoidosis was present in 29/69 patients. AUC – area under the receiver operating characteristic curve, CI – confidence interval, LA – left atrium, LV – left ventricle, RA – right atrium, RV – right ventricle

When evaluating different quantification methods utilizing LA-shrunk as the background (**Figure 3**), CMA had the highest performance followed by VOI (AUC 0.903, 95% CI 0.834 – 0.971), TBR (AUC 0.891, 95% CI 0.819 – 0.964), and SUVmax (AUC 0.812, 95% CI 0.701 – 0.923). Thresholds for each metric, with the associated sensitivity and specificity, are shown in **Table 3**. Abnormal CMA (≥1) had a sensitivity of 100% and specificity 65% for cardiac sarcoidosis. A comparison of thresholds for different background activity are shown in **Supplemental Table 1**. Quantification based on maximal background count or mean background count + 2 standard deviations had the highest prediction performance. A comparison of thresholds for VOI and CMA, with corresponding sensitivity and specificity, is shown in **Supplemental Table 2**.

**Figure 3.**
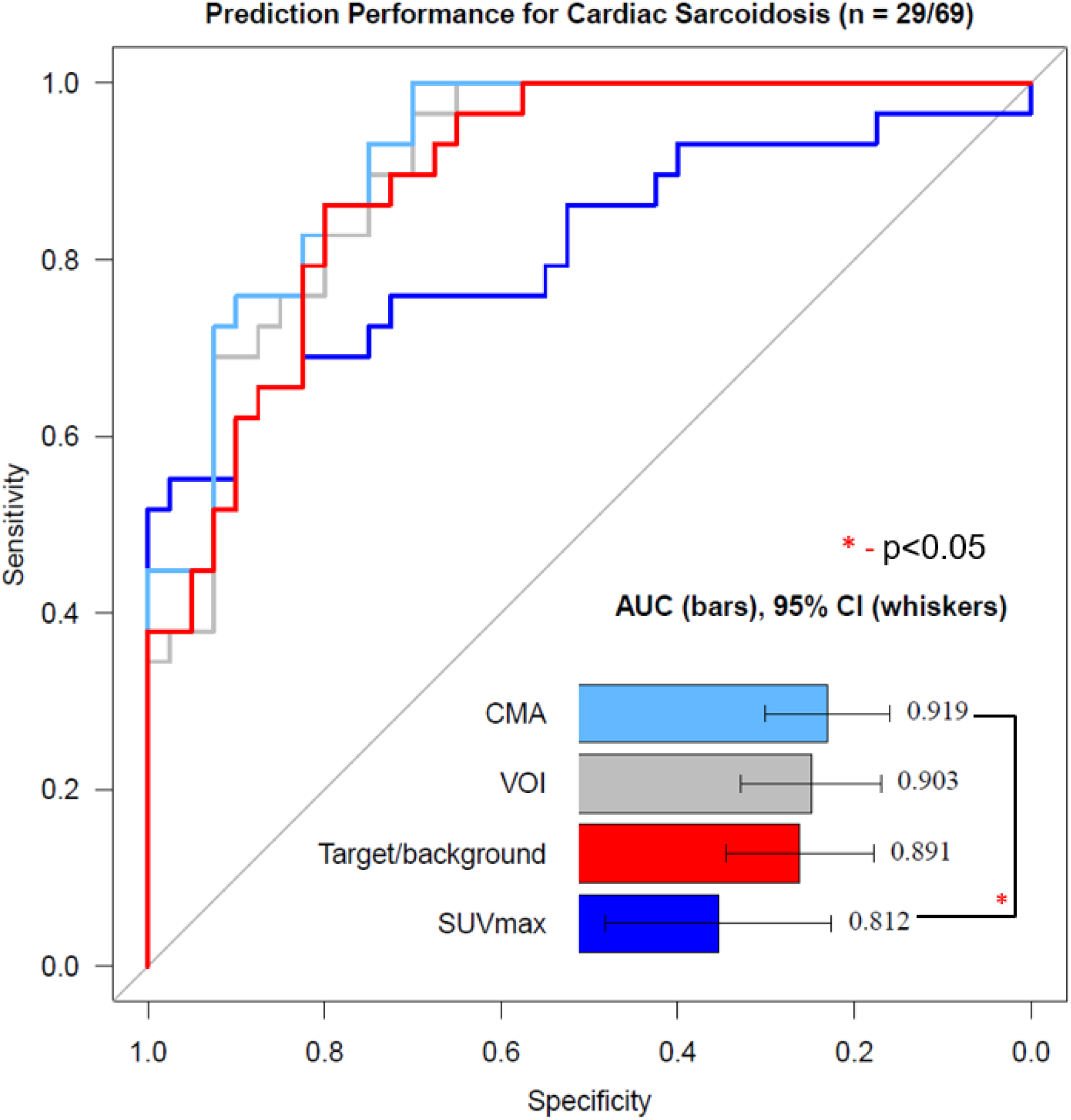
Prediction performance for cardiac sarcoidosis using methods based on a shrunk left atrium background. The area under the receiver operating characteristic curve (AUC) for cardiometabolic activity (CMA), volume of inflammation (VOI), target to background ratio, and maximum standardized uptake value (SUVmax) were not significantly different. CI – confidence interval

**Table 3:** Thresholds for quantitative measures. Sensitivity and specificity are for diagnosis of cardiac sarcoidosis. CMA – cardiometabolic activity, SUVmax – maximum standardized uptake value, VOI – volume of inflammation

|  | Threshold | Sensitivity | Specificity |
| --- | --- | --- | --- |
| SUVmax | $\geq 2.88$ | 66 | 88 |
| Target to Background Ratio | $\geq 1.16$ | 86 | 83 |
| VOI | $> 0$ | 100 | 53 |
| CMA | $\geq 1$ | 100 | 65 |

### Prediction Performance for Pulmonary Abnormalities

Pulmonary abnormalities were present in 34 patients, including 21 with isolated lymphadenopathy and 13 patients with lymphadenopathy in combination with pulmonary nodules or consolidations. Prediction performance for pulmonary abnormalities is shown in **Supplemental Figure 3**. LMA had the highest prediction performance (AUC 0.733, 95% CI 0.613 – 0.852), and had significantly higher AUC compared to LMV (AUC 0.609, 95% CI 0.474 – 0.744, p=0.004).

### Case Examples

An example of quantification in a patient with cardiac sarcoidosis is shown in **Figure 4**. Using LA-shrunk background, the patient had elevated values for SUVmax (9.83), TBR (5.23), VOI (111 mL) and CMA (1052). Diagnosis of cardiac sarcoidosis was established by reduced left ventricular ejection fraction, typical pattern of late gadolinium enhancement, and visually active PET.

**Figure 4.**
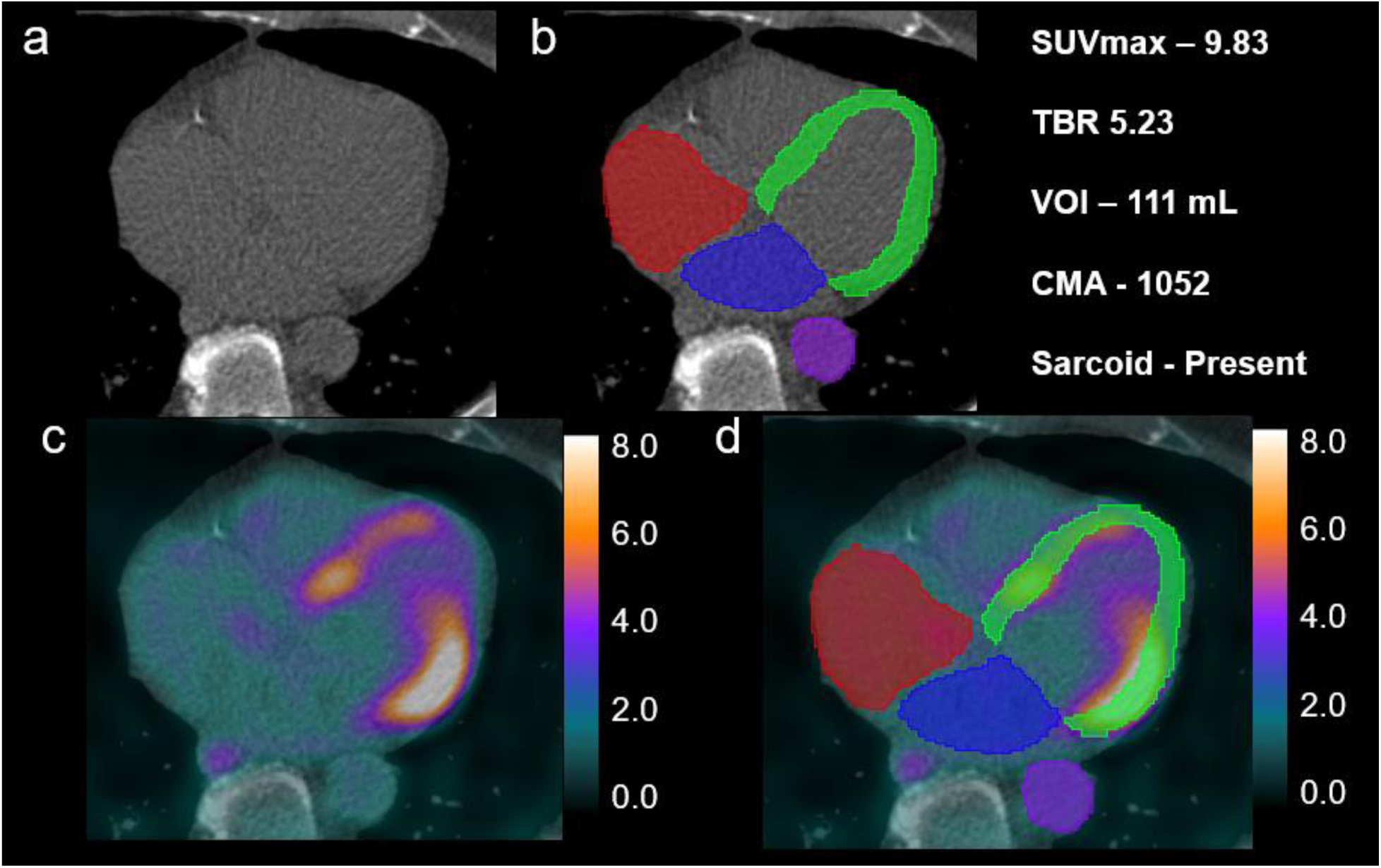
Case example in a man with cardiac sarcoidosis. Computed tomography imaging (Panel a) is segmented using deep learning model (Panel b) to derive left atrial (blue), right atrial (red), aorta (purple) and left ventricular myocardium (green). Fused imaging is shown in Panel c, with scale shown in standardized uptake values (SUV). Chamber segmentations are applied to the co-registered image sets (Panel d) in order to quantify radiotracer activity. The study was positive by visual interpretation and had abnormal quantification. CMA – cardiometabolic activity, SUVmax – maximum SUV, TBR – target to background ratio, VOI – volume of inflammation

**Figure 5** shows quantification in a patient without cardiac sarcoidosis. In the second case, using LA-shrunk background the SUVmax was 1.84, TBR was 0.99, and both CMA and VOI were 0. The patient had no criteria for cardiac sarcoidosis other than reduced left ventricular ejection fraction.

**Figure 5.**
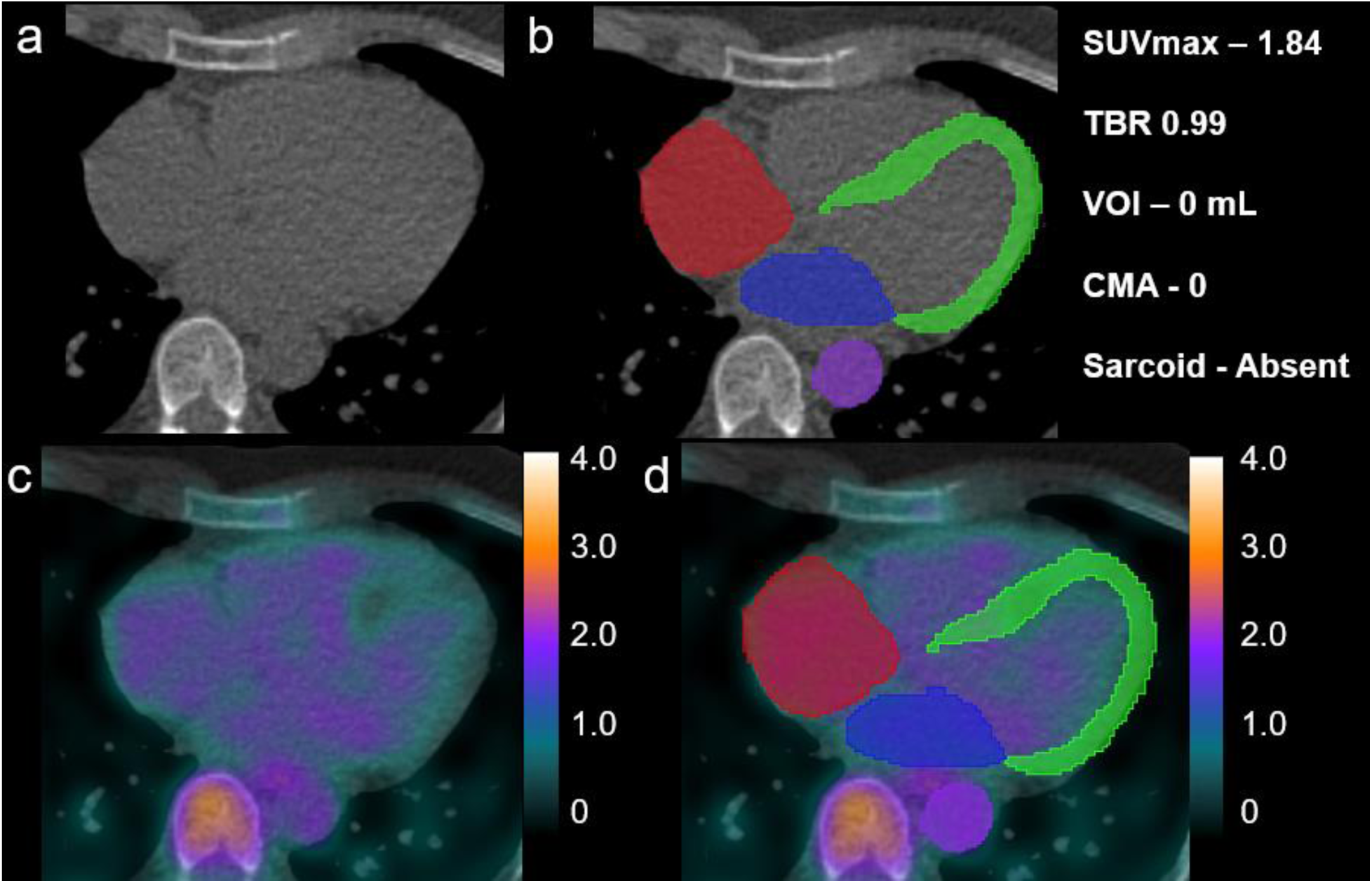
Case example in a man without cardiac sarcoidosis. The patient had a reduced left ventricular ejection fraction but no other criteria for cardiac sarcoidosis. Computed tomography imaging (Panel a) is segmented using deep learning model (Panel b) to derive left atrial (blue), right atrial (red), aorta (purple) and left ventricular myocardium (green). Fused imaging is shown in Panel c, with scale shown in standardized uptake values (SUV). The study was reported as possible uptake in the mid-ventricular septum. Chamber segmentations are applied to the co-registered image sets (Panel d) to quantify radiotracer activity. Deep learning quantification was normal. The patient’s only criteria for sarcoidosis were possible active PET and reduced ventricular function. CMA – cardiometabolic activity, SUVmax – maximum SUV, TBR – target to background ratio, VOI – volume of inflammation

**Figure 6** shows a case with cardiac sarcoidosis with less intense PET activity which was confirmed pathologically in the explanted heart. The patient had mildly elevated values for SUVmax (2.24), TBR (1.32), VOI (4 mL) and CMA (8) using LA-shrunk background. An example of pulmonary quantification is shown in **Supplemental Figure 4**.

**Figure 6.**
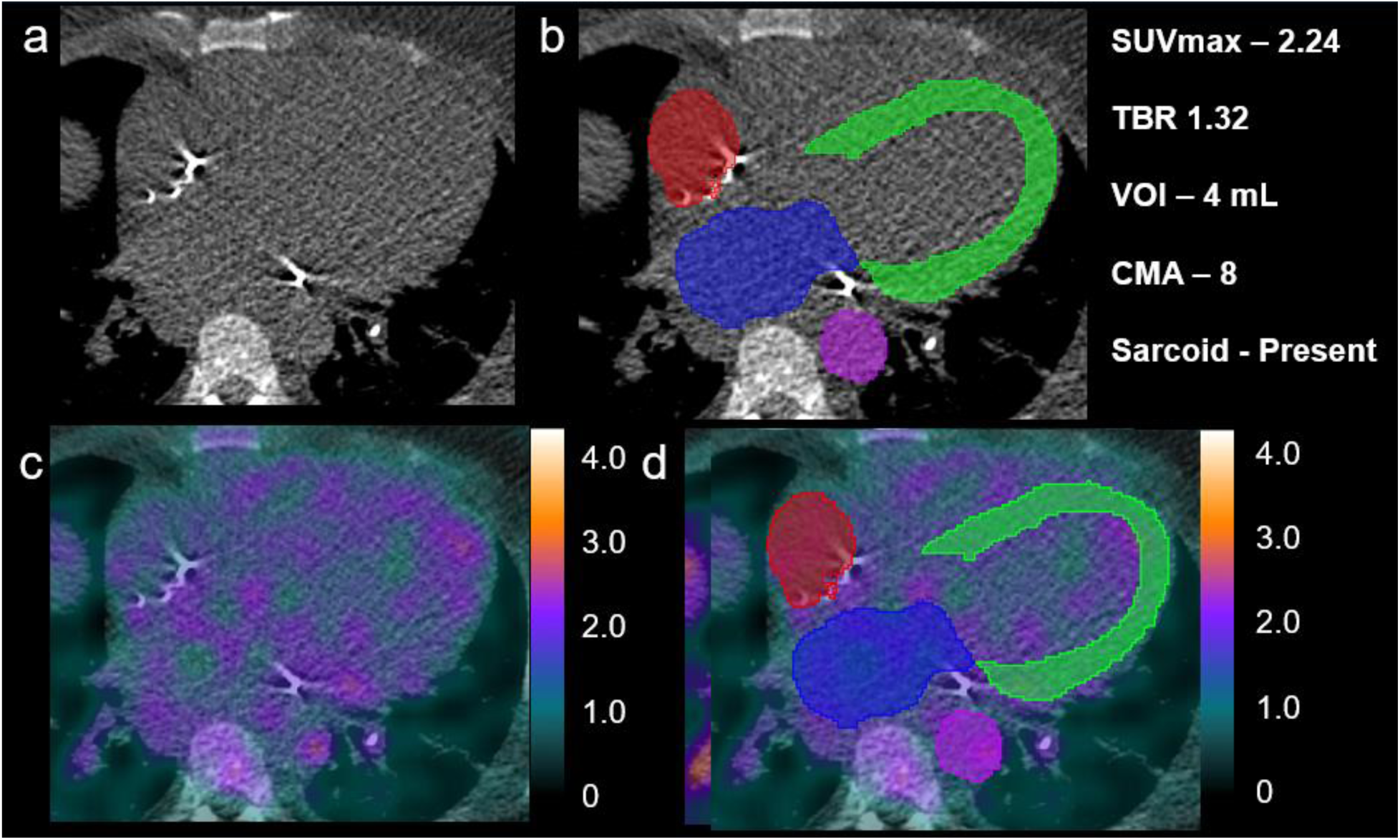
Case example in a man with cardiac sarcoidosis confirmed in the explanted heart. Computed tomography imaging (Panel a) is segmented using deep learning model (Panel b) to derive left atrial (blue), right atrial (red), aorta (purple) and left ventricular myocardium (green). Fused imaging is shown in Panel c, with scale shown in standardized uptake values (SUV). Chamber segmentations are applied to the co-registered image sets (Panel d) in order to quantify radiotracer activity. Deep learning quantification was mildly abnormal, with possible uptake visualized in the distal anterior wall. CMA – cardiometabolic activity, SUVmax – maximum SUV, TBR – target to background ratio, VOI – volume of inflammation

## DISCUSSION

We demonstrated the feasibility of fully automated quantification of [18F]FDG PET by leveraging recent developments in deep learning-based segmentation of CT attenuation maps. Quantification could be performed rapidly, with a mean processing time of ~16 second, without the need for manual definition of myocardial or background regions. Since CT imaging is utilized for the anatomic definition, the quality of the segmentation is not dependent on the pattern or intensity of myocardial uptake, as is the case for conventional approaches. Full automation allowed us to evaluate multiple quantitative metrics simultaneously (including cardiac and pulmonary quantification), with perfect reproducibility since no operator interaction was involved. This approach could be integrated into clinical practice to provide physicians with consistent quantitative metrics of cardiac inflammation to guide diagnosis and treatment recommendations in patients with suspected cardiac sarcoidosis.

Quantification of [18F]FDG PET studies may aid physicians in establishing a diagnosis of cardiac sarcoidosis [9, 11, 19]. This could be important for centers with less expertise or in cases that are equivocal. In our analyses, the accuracy for [18F]FDG quantification obtained by the fully automatic processing was excellent for most of the quantitative metrics. In fact, the prediction performance for CMA (using shrunk LA background and SUVmax threshold) was numerically higher than expert visual interpretation (in spite of the latter being used as part of the diagnostic criteria). Additionally, we demonstrated that our approach could be used to quantify radiotracer activity related to pulmonary abnormalities which may be a helpful aid for interpreting physicians without a radiology background [20]. However, a more clinically relevant application may be improving reproducibility of serial measurements of [18F]FDG uptake. Variations in region of interest placement would be minimized with the proposed approach. Additionally, it enables automated region placement for background activity which improves reproducibility [8]. Previous studies have shown that improvement in quantitative measures of [18F]FDG PET are associated with improvement in ventricular function [21], and may have prognostic importance [22]. Since physicians may use these serial measurements to adjust immunosuppressive therapy [23, 24], it is critically important that they can be performed in a consistent and highly reproducible manner.

In our analysis, abnormal CMA was able to identify patients with cardiac sarcoidosis with a high degree of sensitivity (100%) while maintaining specificity (65%). In contrast, we found that abnormal values for SUVmax were able to identify patients with higher specificity (88%) than sensitivity (66%). Physicians could potentially utilize both measures to maximize both sensitivity and specificity. Additionally, as with any threshold, greater extents of abnormal quantitation should lead physicians to diagnose cardiac sarcoidosis with more confidence. For example, the CMA was much higher in our first presented case (which was interpreted clinically as positive) compared to the third case (which was interpreted clinically as equivocal). The thresholds we identified for maximal background activity are potentially more intuitive clinically compared to thresholds based on mean and standard deviation. The former identifies any measurable activity as abnormal, while with the latter, very small amounts of activity would be considered normal (since ~3% of blood pool activity could fall within the abnormal range).

We evaluated different potential background regions as well as thresholds for abnormal activity. Regarding selection of the background region, the use of shrunk LA, shrunk RV, or shrunk RA backgrounds tended towards higher prediction performance in analyses using maximal background activity as the threshold. However, we also identified a relationship between the threshold for abnormal activity and choice of background region. In particular, we saw less variability in prediction performance by utilizing a threshold based on the mean and standard deviation of background activity and no significant difference in prediction performance using TBR. The two findings in concert suggest that background activity measurements can be influenced by small volumes of adjacent myocardial or lymph node activity. Therefore, quantification using thresholds based on maximal activity seem to benefit from smaller regions of interest to avoid this adjacent hypermetabolism. Meanwhile, utilizing thresholds based on mean activity minimizes the impact on threshold values. These observations are potentially relevant for automated quantification of a variety of hot spot imaging tests.

Our study has a few limitations, the most important of which is that the sample size is limited, allowing us only to demonstrate the feasibility of the approach. However, the fully automated workflow can facilitate larger, multi-center studies in the future. Secondly, we relied on existing clinical criteria for establishing the diagnosis, since cardiac pathology was not available in all patients. However, this reflects current clinical practice and limits bias that may be related to requiring cardiac pathology. Lastly, our method (just like all hybrid imaging) relies on accurate registration between image sets, and this needs to be assessed by technologists during the reconstruction process.

## CONCLUSIONS

We demonstrate that fully automated quantification of [18F]FDG PET for cardiac sarcoidosis, utilizing chamber segmentation from the CT attenuation maps, is feasible and has high diagnostic accuracy. This approach could be applied to provide objective and reproducible measurements of cardiac hypermetabolism to aid in diagnosis or follow response to therapy.

## SOURCES OF FUNDING

This research was supported in part by grant R35HL161195 from the National Heart, Lung, and Blood Institute/ National Institutes of Health (NHLBI/NIH) (PI: Piotr Slomka). The content is solely the responsibility of the authors and does not necessarily represent the official views of the National Institutes of Health.

## DISCLOSURES

Dr. Miller reports research and consulting support from Pfizer. Mr. Kavanagh, Drs. Berman and Slomka participate in software royalties for QPS software at Cedars-Sinai Medical Center. Dr. Slomka has received research grant support from Siemens Medical Systems and consulting fees from Synektik. Dr. Berman has served as a consultant for GE Healthcare. Dr. Hidesato Fujito received research funding support from the Fukuda Foundation for Medical Technology, the Nihon University School of Medicine Alumni Association Research Grant and the Society of Nuclear Medicine and Molecular Imaging Wagner-Torizuka Fellowship grant. The authors have no other relevant disclosures.

## Data Availability

All data produced in the present study are available upon reasonable request to the authors.

## Notes

### Author Declarations

This study was approved by the institutional review board at Cedars-Sinai Medical Center including waiver of informed consent for use of retrospective data.

