## Supplemental Material for "AI-enabled CT-guided end-to-end quantification of total cardiac activity in 18FDG cardiac PET/CT for detection of cardiac sarcoidosis"

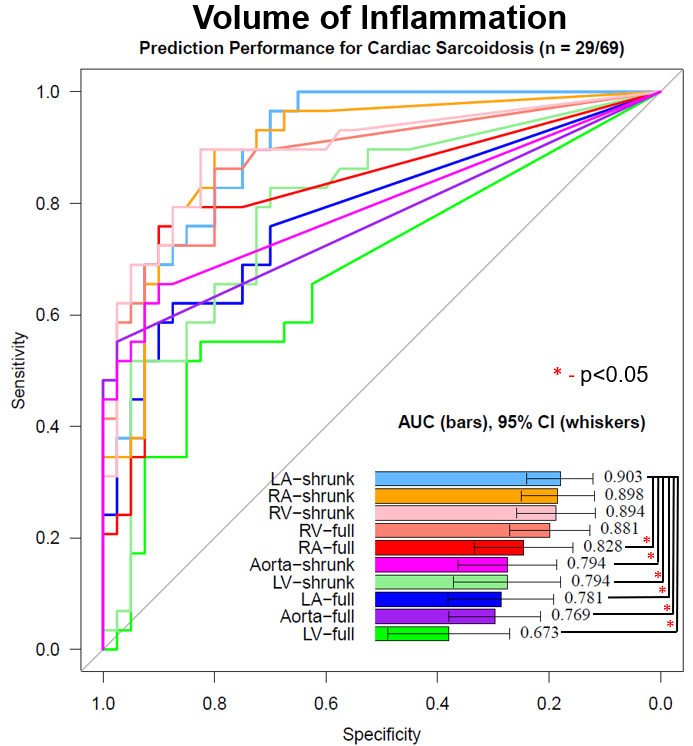

**Supplemental Fig. 1** Prediction performance for cardiac sarcoidosis using different methods for quantifying volume of inflammation. Segments were derived using deep learning from computed tomography attenuation imaging. AUC – area under the receiver operating characteristic curve, CI – confidence interval, LA – left atrium, LV – left ventricle, RA – right atrium, RV – right ventricle.

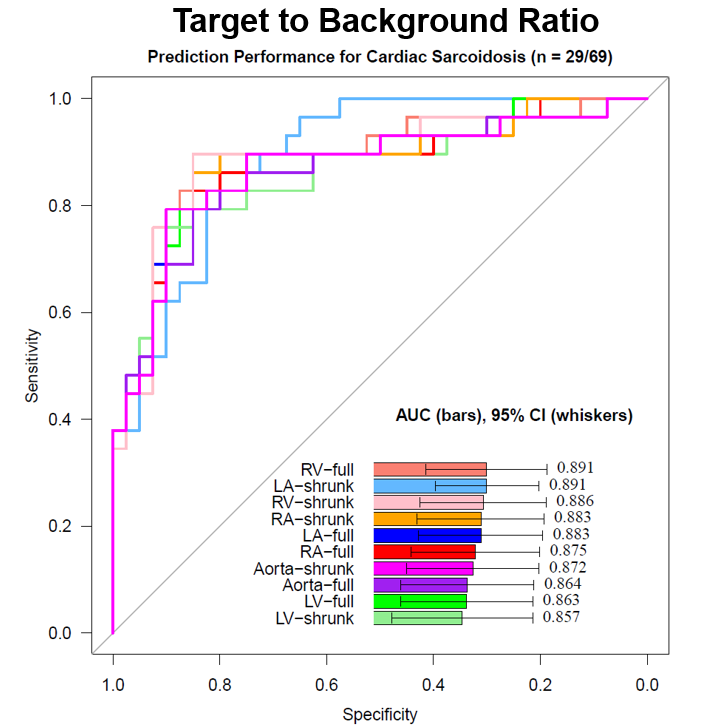

**Supplemental Fig. 2** Prediction performance for cardiac sarcoidosis using different methods for quantifying target to mean background ratio. Segments were derived using deep learning from computed tomography attenuation imaging. AUC – area under the receiver operating characteristic curve, CI – confidence interval, LA – left atrium, LV – left ventricle, RA – right atrium, RV – right ventricle

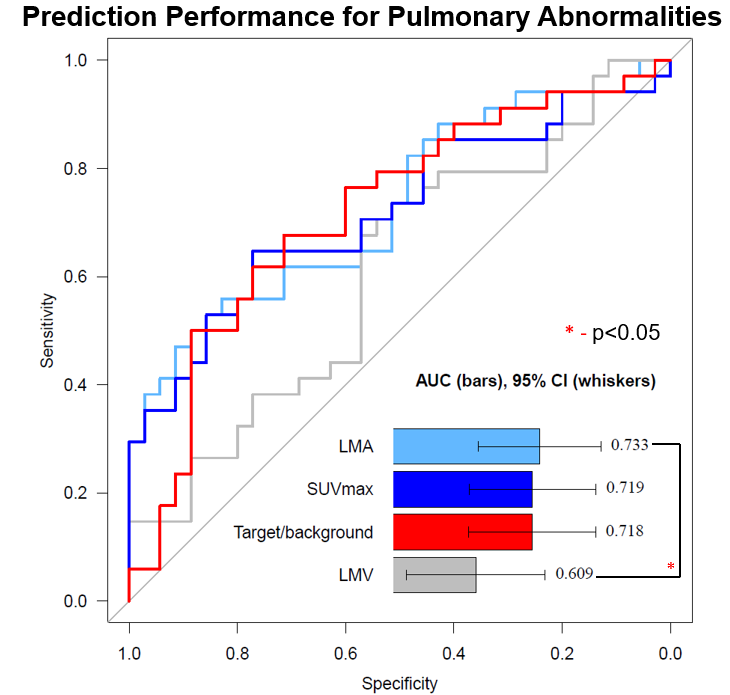

**Supplemental Fig. 3** Prediction performance for pulmonary abnormalities (N=34/69) using different methods for quantifying lung radiotracer activity. Pulmonary abnormalities included lymphadenopathy, pulmonary nodules, or consolidations. Segments were derived using deep learning from computed tomography attenuation imaging. AUC – area under the receiver operating characteristic curve, CI – confidence interval, LMA – lung metabolic activity, LMV – lung metabolic volume, SUVmax – maximum standardized uptake value

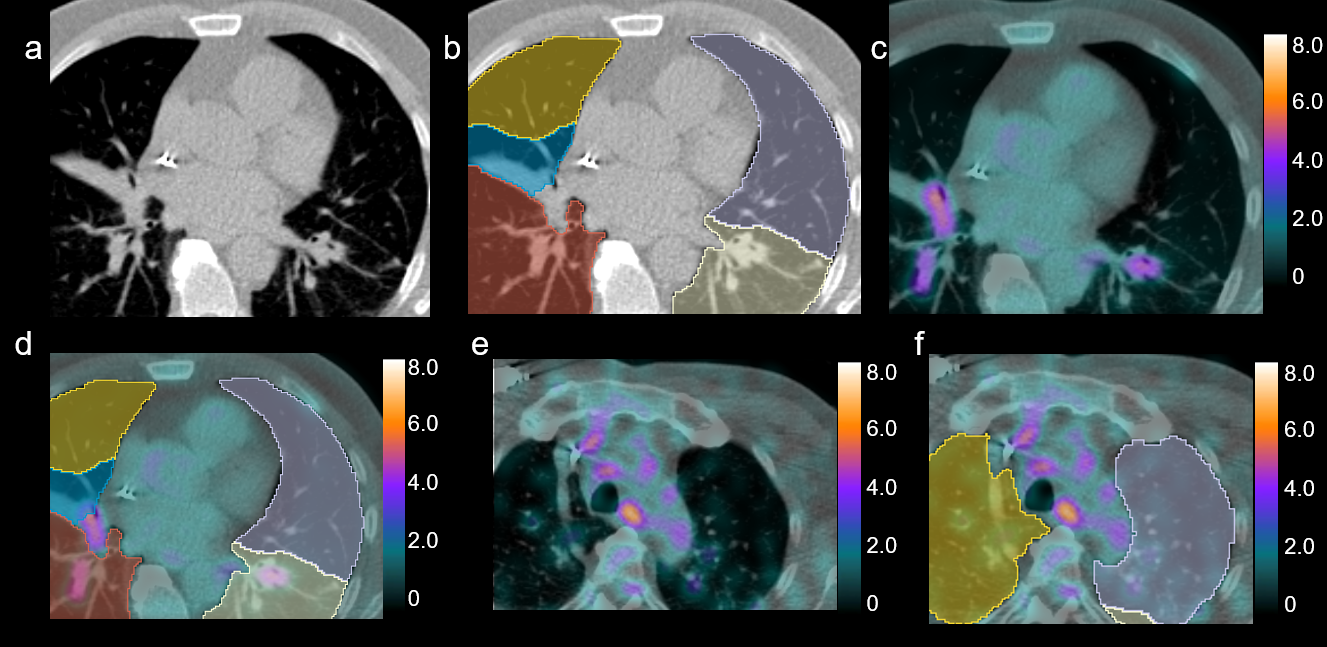

**Supplemental Fig. 4** Example of pulmonary quantification. Computed tomography attenuation correction scans (panel a) were segmented to identify lung segments shown in panel b (left lower lobe - white, left upper lobe - lavender, right upper lobe - yellow, right middle lobe - blue, right lower lobe - red). Fused imaging (panel c and e) can be quantified using the same segmentation to quantify uptake related to lymph nodes (left and right lower lobes – panel d) and parenchymal abnormalities such as pulmonary nodules (left upper lobe panel f). The maximal standardized uptake value was 3.8, with target to mean background ratio of 16, lung metabolic volume of 110mL and lung metabolic activity of 136.

|  | Max Background | Max Background *1.3 | Max Background *1.5 | Mean background + 2 SD |
| --- | --- | --- | --- | --- |
| Volume of inflammation |  |  |  |  |
| Full left atrium | 0.781 (0.672 – 0.890) | 0.691 (0.586 – 0.795) | 0.678 (0.585 – 0.772) | **0.908 (0.841 – 0.975)** |
| Shrunk left atrium | 0.903 (0.834 – 0.971) | 0.776 (0.675 – 0.877) | 0.731 (0.630 – 0.832) | **0.916 (0.852 – 0.980)** |
| Full left ventricle | 0.673 (0.548 – 0.799) | 0.524 (0.459 – 0.599) | 0.491 (0.444 – 0.539) | **0.867 (0.783 – 0.951)** |
| Shrunk left ventricle | 0.794 (0.683 – 0.904) | 0.693 (0.594 – 0.792) | 0.597 (0.510 – 0.683) | **0.884 (0.807 – 0.961)** |
| Full right atrium | 0.828 (0.726 – 0.929) | 0.722 (0.618 – 0.826) | 0.703 (0.605 – 0.802) | **0.881 (0.795 – 0.967)** |
| Shrunk right atrium | **0.898 (0.823 – 0.973)** | 0.784 (0.685 – 0.884) | 0.712 (0.610 – 0.813) | 0.884 (0.801 – 0.966) |
| Full right ventricle | 0.882 (0.799 – 0.964) | 0.655 (0.569 – 0.741) | 0.586 (0.516 – 0.656) | **0.883 (0.794 – 0.971)** |
| Shrunk right ventricle | **0.894 (0.813 – 0.975)** | 0.775 (0.675 – 0.874) | 0.663 (0.571 – 0.754) | 0.883 (0.798 – 0.968) |
| Full aorta | 0.769 (0.674 – 0.863) | 0.690 (0.600 – 0.780) | 0.655 (0.569 – 0.741) | **0.876 (0.793 – 0.959)** |
| Shrunk aorta | 0.794 (0.693 – 0.895) | 0.728 (0.630 – 0.827) | 0.724 (0.632 – 0.816) | **0.874 (0.788 – 0.960)** |
| Cardiometabolic Activity |  |  |  |  |
| Full left atrium | 0.787 (0.679 – 0.895) | 0.692 (0.588 – 0.796) | 0.681 (0.588 – 0.774) | **0.916 (0.853 – 0.978)** |
| Shrunk left atrium | **0.919 (0.858 – 0.980)** | 0.779 (0.679 – 0.880) | 0.734 (0.633 – 0.835) | 0.909 (0.841 – 0.976) |
| Full left ventricle | 0.688 (0.564 – 0.812) | 0.524 (0.459 – 0.590) | 0.492 (0.444 – 0.540) | **0.879 (0.801 – 0.957)** |
| Shrunk left ventricle | 0.805 (0.699 – 0.911) | 0.696 (0.597 – 0.795) | 0.597 (0.510 – 0.683) | **0.891 (0.818 – 0.965)** |
| Full right atrium | 0.837 (0.739 – 0.936) | 0.725 (0.621 – 0.829) | 0.703 (0.605 – 0.802) | **0.885 (0.801 – 0.970)** |
| Shrunk right atrium | **0.904 (0.832 – 0.976)** | 0.789 (0.690 – 0.888) | 0.713 (0.611 – 0.814) | 0.893 (0.814 – 0.972) |
| Full right ventricle | 0.893 (0.813 – 0.973) | 0.655 (0.569 – 0.741) | 0.586 (0.516 – 0.656) | **0.894 (0.809 – 0.979)** |
| Shrunk right ventricle | **0.901 (0.823 – 0.979)** | 0.779 (0.680 – 0.877) | 0.663 (0.572 – 0.755) | 0.893 (0.811 – 0.976) |
| Full aorta | 0.752 (0.656 – 0.847) | 0.690 (0.600 – 0.780) | 0.655 (0.569 – 0.741) | **0.882 (0.801– 0.962)** |
| Shrunk aorta | 0.796 (0.695 – 0.896) | 0.728 (0.630 – 0.827) | 0.724 (0.632 – 0.816) | **0.878 (0.793 – 0.962)** |

**Supplemental Table 1.** Prediction performance for cardiac sarcoidosis using different thresholds for background activity. Highest prediction performance for each background region shown in bold. SD – standard deviation.

|  | Max Background | | | Mean background + 2 SD | | |
| --- | --- | --- | --- | --- | --- | --- |
|  | Threshold | Sensitivity | Specificity | Threshold | Sensitivity | Specificity |
| Volume of inflammation |  |  |  |  |  |  |
| Full left atrium | 5 | 62 | 88 | 6 | 86 | 85 |
| Shrunk left atrium | 0 | 97 | 70 | 11 | 86 | 88 |
| Full left ventricle | 0 | 55 | 82 | 6 | 72 | 88 |
| Shrunk left ventricle | 0 | 83 | 70 | 5 | 79 | 82 |
| Full right atrium | 1 | 79 | 88 | 2 | 83 | 85 |
| Shrunk right atrium | 1 | 76 | 93 | 2 | 83 | 88 |
| Full right ventricle | 0 | 86 | 80 | 2 | 83 | 90 |
| Shrunk right ventricle | 0 | 90 | 82 | 2 | 83 | 88 |
| Full aorta | 0 | 52 | 97 | 0 | 86 | 85 |
| Shrunk aorta | 0 | 66 | 90 | 0 | 86 | 85 |
| Cardiometabolic Activity |  |  |  |  |  |  |
| Full left atrium | 12 | 62 | 90 | 11 | 86 | 82 |
| Shrunk left atrium | 1 | 100 | 65 | 18 | 86 | 85 |
| Full left ventricle | 1 | 55 | 85 | 15 | 72 | 85 |
| Shrunk left ventricle | 1 | 83 | 70 | 11 | 79 | 82 |
| Full right atrium | 10 | 76 | 93 | 8 | 83 | 85 |
| Shrunk right atrium | 20 | 76 | 93 | 12 | 83 | 88 |
| Full right ventricle | 0 | 90 | 80 | 10 | 83 | 90 |
| Shrunk right ventricle | 7 | 79 | 90 | 12 | 83 | 90 |
| Full aorta | 0 | 52 | 97 | 0 | 86 | 85 |
| Shrunk aorta | 1 | 66 | 90 | 1 | 86 | 85 |

**Supplemental Table 2.** Comparison of thresholds for volumetric measurements of hypermetabolism. SD – standard deviation.
